# Wearable-Derived Long-Term Behavioral Patterns and Short-Term Dynamics Associated With Depressive Symptom Severity

**DOI:** 10.64898/2026.05.27.26354070

**Authors:** Jenifer Rim, Qi Xu, Xiwei Tang, Cadence Pinkerton, Yuqing Guo, Annie Qu

**Affiliations:** Department of Statistics, University of California, Irvine, CA, USA; Department of Statistics and Data Science, Carnegie Mellon University, Pittsburgh, PA, USA; Department of Mathematical Sciences, University of Texas at Dallas, Richardson, TX, USA; Sue & Bill Gross School of Nursing, University of California, Irvine, CA, USA; Department of Statistics and Applied Probability, University of California, Santa Barbara, CA, USA

**Keywords:** depression, wearable, Fitbit, physical activity, sleep, digital phenotyping, PHQ-9

## Abstract

**Background:** Wearable-based studies have largely examined activity and sleep using static summaries or single time windows, potentially missing how chronic patterns and recent behavioral changes jointly relate to depressive symptom severity. We evaluated whether combining long-term habitual behavior with short-term dynamics improves characterization of moderate-to-severe depressive symptoms.

**Methods:** We analyzed Fitbit data from All of Us participants with Patient Health Questionnaire-9 (PHQ-9) assessments, defining moderate-to-severe symptoms as PHQ-9 ≥ 10 (*N* = 248). Logistic regression evaluated long-term measures (past-year step count and awake time after sleep onset) and short-term dynamics (30-day step decline and 30-day sleep duration variability), adjusting for demographics. Performance was assessed via repeated stratified 10-fold cross-validation.

**Results:** Thirty percent of participants (*n* = 74) had moderate-to-severe depressive symptoms. Higher long-term step count was associated with lower odds of elevated symptoms (OR = 0.75 per 1,000 steps/day), greater awake time after sleep onset with higher odds (OR = 1.27 per 1%), a 30-day step decline with higher odds (OR = 2.70), and greater 30-day sleep variability with higher odds (OR = 1.07 per percentage point). Short-term dynamics provided complementary information beyond long-term measures alone. The combined model achieved the highest discrimination (area under the curve [AUC] = 0.80 vs. 0.73 demographics-only), though findings should be interpreted as exploratory given the modest sample size.

**Limitations:** The sample was modest in size (*N* = 248), PHQ-9 reflects symptom severity rather than clinical diagnosis, causal inference is not possible given the cross-sectional outcome assessment, and Fitbit users may not represent broader populations.

**Conclusions:** Long-term behavioral patterns and short-term changes in activity and sleep were associated with depressive symptom severity, supporting wearable-derived measures as potential adjunctive markers in mental health research.

**Highlights:**

- Higher long-term step count was associated with lower odds of depression
- A 30-day step decline was linked to 170% higher odds of elevated symptoms
- Greater 30-day sleep variability was associated with depressive symptoms
- Combined long- and short-term features improved discrimination (AUC = 0.80)
- Wearable behavioral dynamics may serve as adjunctive markers in mental health research

## 1 Introduction

Depression is one of the leading causes of disability worldwide, contributing substantially to reduced quality of life, lost productivity, and increased healthcare costs (Friedrich, 2017). In the United States, nearly one in five adults will experience major depressive disorder during their lifetime, with moderate-to-severe forms associated with greater functional impairment and comorbidity (Lee et al., 2023; Kessler et al., 2003). Beyond affective symptoms, depression is also associated with disruptions in motivation, energy, psychomotor activity, sleep, and daily functioning (American Psychiatric Association, 2013). These changes often emerge gradually and fluctuate over time before individuals seek clinical care, highlighting the need for scalable approaches capable of capturing patterns related to depressive symptom severity outside of traditional clinical settings (Smith et al., 2013).

Wearable devices such as Fitbit have emerged as promising tools for continuously measuring physical activity, sleep, and other health-related measures in free-living environments (Wang et al., 2022). By passively collecting data over extended periods, wearable devices may capture fluctuations in daily functioning that are difficult to assess through episodic surveys or clinical evaluations (Moshe et al., 2021). This approach aligns with the broader framework of digital phenotyping, which uses data from personal digital devices to characterize individual-level patterns in naturalistic settings (Torous et al., 2016; Insel, 2017). Passive sensing approaches may therefore provide objective wearable-derived markers associated with depressive symptom severity while reducing participant burden and improving ecological validity (Jacobson and Chung, 2020).

Prior studies have demonstrated that lower physical activity and disrupted sleep are associated with greater depressive symptom severity (Lim et al., 2022; Bizzozero-Peroni et al., 2024; Maki et al., 2025). Objective measures such as daily step counts and sleep patterns have been linked to both incident depression and symptom severity across diverse populations, with associations thought to reflect fatigue, reduced motivation, psychomotor slowing, and withdrawal from daily activities (Bizzozero-Peroni et al., 2024; Maki et al., 2025). Large-scale meta-analyses further support a dose-response relationship between physical activity and depression risk, suggesting that even modest increases in activity may confer protective benefits (Pearce et al., 2022). Sleep disturbances, including irregular duration and reduced sleep efficiency, have also been associated with both the onset and severity of depressive symptoms (Lim et al., 2022; Maki et al., 2025). Emerging evidence further suggests that sleep regularity, beyond duration alone, may represent an important marker of emotional functioning and mental health (Baglioni et al., 2011; Lunsford-Avery et al., 2018).

Although prior wearable-based studies have linked reduced physical activity and disturbed sleep with depression, many studies rely primarily on static summaries averaged across broad time windows or evaluate behavioral measures at a single temporal scale. Such approaches may obscure potentially important distinctions between chronic behavioral patterns and recent within-person behavioral changes preceding elevated depressive symptoms. For example, persistently low activity may reflect long-term behavioral vulnerability, whereas recent declines in activity or increasing sleep irregularity may capture more acute behavioral shifts associated with worsening symptoms. Relatively few studies have jointly evaluated these complementary temporal dynamics using objectively measured wearable data in large population-based cohorts (Snippe et al., 2016; Lindwall et al., 2014; Vidal Bustamante et al., 2020).

From a clinical perspective, temporal behavioral dynamics may provide information not captured by static behavioral averages alone. Whereas long-term habitual activity and sleep patterns may reflect baseline behavioral vulnerability, acute within-person declines in activity or sleep regularity may more closely correspond to worsening depressive symptom severity or emerging functional impairment. These short-term behavioral changes may be particularly important because depressive symptoms often fluctuate between clinical encounters and may not be recognized until substantial deterioration has occurred (Smith et al., 2013). Continuous wearable monitoring therefore offers a unique opportunity to characterize behavioral trajectories in real-world settings and may enable identification of dynamic behavioral changes associated with worsening depressive symptoms beyond what can be inferred from cross-sectional self-report measures alone.

The All of Us Research Program provides a unique opportunity to address these gaps by linking longitudinal wearable data with electronic health records and validated mental health measures across a large and diverse cohort (All of Us Research Program Investigators, 2019). This enables evaluation of wearable-derived markers associated with depressive symptoms in populations historically underrepresented in biomedical research. Prior work has documented racial and ethnic disparities in depression prevalence, treatment access, and diagnosis, driven in part by structural and socioeconomic factors (Williams and Mohammed, 2009; Gonzalez et al., 2010). Leveraging this resource therefore enables evaluation of activity, sleep, and temporal dynamics within a more diverse cohort than many prior wearable-based studies.

In this study, we examine wearable-derived physical activity and sleep measures in relation to depressive symptom severity, moving beyond static summaries by characterizing complementary temporal dynamics. We hypothesize that recent declines in step counts and greater variability in sleep duration are associated with increased odds of moderate-to-severe depressive symptoms, assessed using the Patient Health Questionnaire-9 (PHQ-9, scores ≥ 10) (Kroenke et al., 2001). We further evaluate whether combining long-term habitual patterns with short-term behavioral dynamics provides complementary information regarding depressive symptom severity beyond demographic factors alone. Identifying passively collected behavioral markers associated with worsening depressive symptoms may support future efforts aimed at behavioral monitoring between clinical encounters.

## 2 Methods

### 2.1 Study design and participants

This study was a retrospective observational analysis using longitudinal wearable data from the All of Us Research Program, which is a large, nationwide cohort created by the National Institutes of Health (NIH) in the United States in 2018 to support precision medicine by integrating electronic health records (EHRs), survey responses, biospecimens, and wearable data (All of Us Research Program Investigators, 2019). We used the Registered Tier Dataset version 8, which contains participant data with a cutoff date of October 1, 2023. Participants were eligible if they had both Fitbit data and Patient Health Questionnaire-9 (PHQ-9) survey responses for their depressive symptoms. Individuals were excluded if they had missing demographic or Fitbit data, insufficient Fitbit records, or if their Fitbit data occurred only after the PHQ-9 survey. The final sample included *N* = 248 participants (see Figure 1 for details).

**Figure 1.**
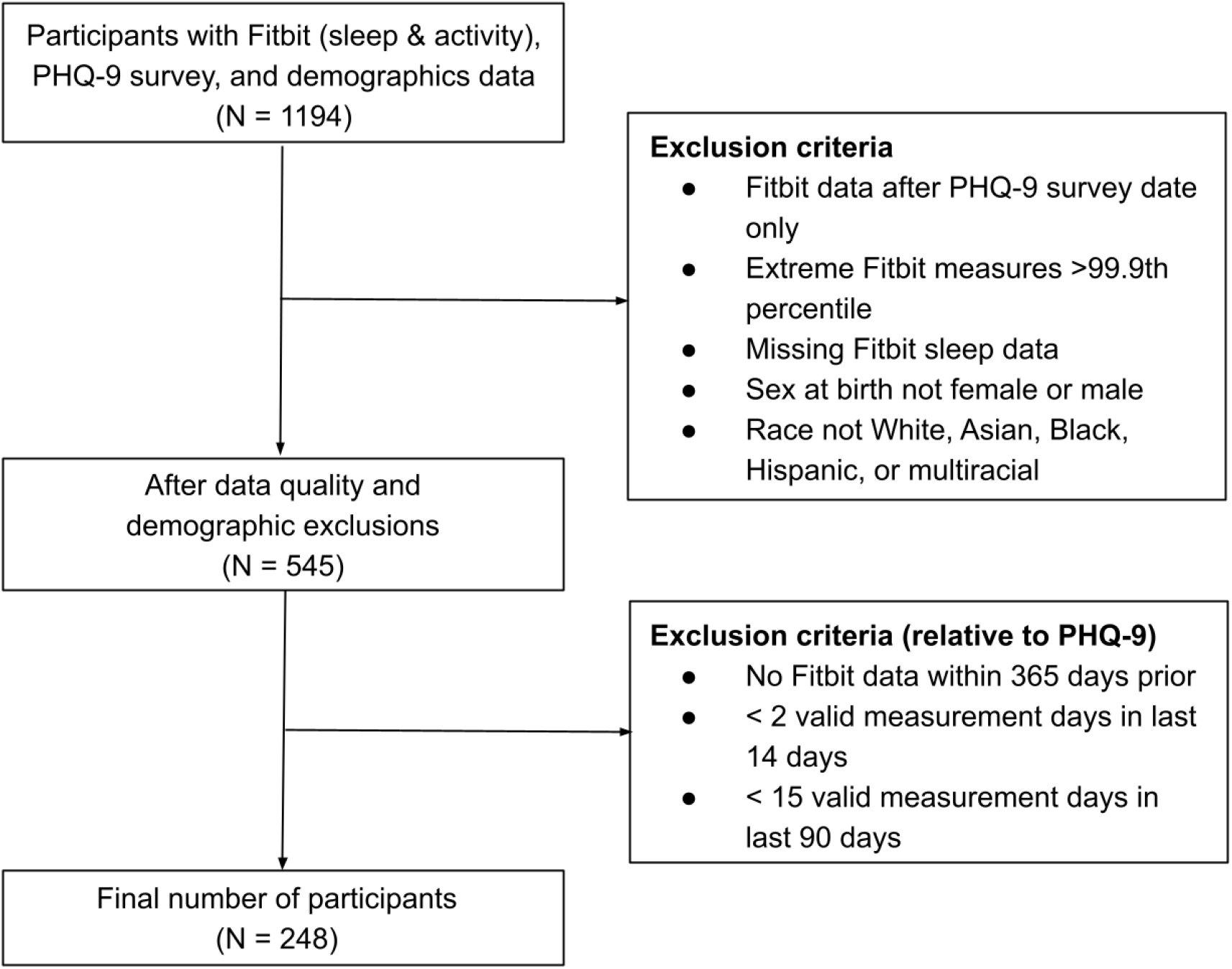
Flowchart of participant inclusion and exclusion criteria.

### 2.2 PHQ-9: depressive symptom assessment

Depressive symptoms were assessed using the Patient Health Questionnaire-9 (PHQ-9), a widely validated self-report instrument designed to screen for, monitor, and measure the severity of depressive symptoms (Kroenke et al., 2001). The PHQ-9 consists of nine items, each reflecting one of the core diagnostic criteria for major depressive disorder, such as depressed mood, loss of interest, sleep disturbance, fatigue, and difficulty concentrating.

Respondents rate the frequency of each symptom over the past two weeks on a 4-point Likert scale ranging from 0 (“not at all”) to 3 (“nearly every day”), yielding a total score between 0 and 27. Higher scores reflect greater depressive symptom severity. Following established cutoffs, we classified scores of 10 or higher as indicating moderate-to-severe depression, while scores below 10 were categorized as no or mild depression (Manea et al., 2012). To ensure comparability across participants, we used the PHQ-9 survey date as an anchor point and aligned all wearable-derived features (e.g., sleep, physical activity) such that they preceded the survey date. This temporal alignment allowed us to examine how both recent and long-term behavioral patterns related to depressive symptom severity, while maintaining consistency in the observation window across individuals.

### 2.3 Fitbit measures and feature engineering

We utilized Fitbit data to obtain objective measures of daily physical activity and sleep, with particular emphasis on behavioral patterns relevant to depressive symptomatology. All time series were temporally aligned relative to each participant’s PHQ-9 survey date, which served as the index date for depression assessment. For each individual, we extracted a 30-day window of Fitbit data preceding the survey date to capture recent physical activity and sleep behavior. A valid Fitbit day was defined as one with more than 1,000 recorded steps (Carrasco et al., 2019) to ensure inclusion of days with sufficient wear time and reliable activity tracking.

Feature construction was guided by the need to capture both habitual long-term behavioral patterns (past year from survey date) and acute short-term fluctuations (past 30 days from survey date) that may be related to depressive symptom severity. For long-term habitual behavior, we derived the average daily step count as a measure of overall physical activity and the average proportion of time spent awake after sleep onset (“average awake %”) as an indicator of sleep fragmentation and poor sleep continuity. These features provide a summary of habitual lifestyle patterns related to depressive symptom severity.

To characterize short-term behavioral changes, we computed trajectory-based features over the 30 days preceding the PHQ-9 survey. Specifically, for each participant *i*, let *s*_*it*_ denote the step count on day *t*, where *t* ∈ {−30, …, 0} indexes valid Fitbit days before the PHQ-9 survey date (*t* = 0). We fit a simple linear regression model,

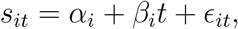

and obtained the ordinary least squares (OLS) slope estimate 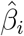. A binary indicator of declining physical activity was then defined as

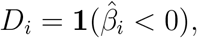

which was equal to 1 if the participant’s step count demonstrated a net downward trend over the 30-day window and 0 otherwise. This binary indicator, rather than the continuous slope estimate, was included in the logistic regression model. A binary indicator was selected to improve interpretability and reduce sensitivity to noisy day-to-day fluctuations in individual slope estimates.

For sleep, we quantified irregularity in sleep duration using the coefficient of variation (CV) rather than the standard deviation, as CV normalizes variability relative to an individual’s average sleep duration and therefore facilitates comparison across participants with different baseline sleep levels. Specifically, the CV for participant *i* was defined as

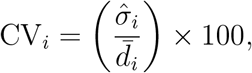

where 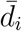 represents the average total sleep duration in minutes and 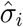 represents the standard deviation of total sleep duration across valid days within the 30-day window. Multiplying by 100 expresses the CV as a percentage, such that a one-unit increase corresponds to a 1 percentage point increase in relative sleep variability.

These Fitbit-derived measures, including past-year average step count, past-year average awake %, 30-day decline in step count, and 30-day CV of sleep duration, were summarized at the participant level and included in subsequent analyses to evaluate their associations with moderate-to-severe depressive symptoms alongside demographic covariates including age, sex, and race. Race categories included White (n = 184), Multiracial (n = 29), Hispanic (n = 15), Asian (n = 12), and Black (n = 8). Due to limited subgroup sample sizes and class imbalance, racial categories were collapsed into White versus non-White groups to improve statistical stability and reduce sparse-data bias. This categorization was used solely for analytic purposes and should not be interpreted as implying homogeneity across racial or ethnic groups.

### 2.4 Statistical analysis

We used logistic regression models to assess associations between demographic characteristics, physical activity, sleep features, and the odds of moderate-to-severe depressive symptoms. Models produced odds ratios (ORs) with 95% confidence intervals (CIs) to quantify the relationship between each predictor and depressive symptom status. All models were adjusted for baseline demographic covariates, including age, sex, and race. Coefficient estimates and standard errors reported in Table 2 were obtained by fitting each model on the full sample of 248 participants.

To evaluate potential multicollinearity among predictors, we calculated variance inflation factors (VIFs) and confirmed that all variables were below the commonly used threshold of 5, suggesting that multicollinearity was not a concern (O’Brien, 2007). We also considered the recommended events-per-variable (EPV) guideline of at least 10 observed outcome events per predictor (Peduzzi et al., 1996). With 74 outcome-positive participants and 7 predictors (including 3 demographic covariates), the observed EPV of approximately 10.6 was near but marginally above this threshold; coefficient estimates and confidence intervals should therefore be interpreted with appropriate caution (Peduzzi et al., 1996).

Fitbit-derived measures included multiple physical activity and sleep-related features. For each participant, features were computed across both long-term (e.g., past year) and short-term time windows (e.g., 30, 30–60, and 60–90 days prior to PHQ-9 assessment), which captured average values, temporal trends (slopes), and variability (CV). The 30-day window immediately preceding the survey date was selected to represent acute behavioral changes, whereas the past year captured habitual behavioral patterns. Candidate features were selected a priori based on existing literature and retained based on interpretability and consistency of association in preliminary models.

Out-of-sample classification performance was evaluated using the area under the receiver operating characteristic curve (AUC) (Bradley, 1997), estimated via repeated stratified 10-fold cross-validation with 20 repeats (Kohavi, 1995; Arlot and Celisse, 2010). With only 74 outcome-positive participants, a single held-out test set would contain too few positive cases for stable AUC estimation. Repeated cross-validation addresses this limitation by averaging performance across 200 held-out folds, yielding average AUC ± SD as a more stable estimate of generalization performance. Folds were assigned at the subject level with stratification on outcome status to preserve the event rate across splits. To prevent data leakage, all wearable-derived features were recomputed independently within each training fold so that no information from test-fold participants influenced feature construction. Out-of-fold predictions from the first repeat, in which each subject was predicted exactly once by a model that had not previously observed them, were used to construct the ROC curves shown in Figure 4.

As sensitivity analyses, we additionally modeled PHQ-9 score as a continuous outcome using linear regression to evaluate whether associations observed using the dichotomized PHQ-9 ≥ 10 threshold were directionally consistent across the full range of depressive symptom severity. We also replaced the binary indicator of declining step count with the continuous ordinary least squares slope estimate to assess whether findings were robust to alternative parameterizations of short-term step trajectories.

## 3 Results

### 3.1 Participant characteristics

Table 1 summarizes subject-level demographic characteristics and habitual physical activity and sleep features by depressive symptom status (moderate-to-severe vs. none-to-mild). All wearable-derived measures were collapsed to the subject level prior to analysis. A total of 248 participants were included, of whom 174 (70%) reported none-to-mild depressive symptoms and 74 (30%) reported moderate-to-severe symptoms based on PHQ-9 scores. Average PHQ-9 scores differed markedly between groups (3.38 vs. 16.03), consistent with the classification threshold of ≥ 10.

**Table 1:** Summary of participant characteristics stratified by depressive symptom severity (none-to-mild vs. moderate-to-severe). Continuous variables are summarized as mean (SD), and categorical variables as count (%). *P*-values were obtained from two-sample t-tests using subject-level collapsed measures, and statistically significant values are indicated with an asterisk (*).

| **Characteristic** | **None-to-Mild**<br>n = 174 (70%) | **Moderate-to-Severe**<br>n = 74 (30%) | **$P$-value** |
| --- | --- | --- | --- |
| PHQ-9 score | 3.38 (3.51) | 16.03 (4.50) | — |
| Valid Fitbit days | 182.47 (101.85) | 167.96 (107.56) | — |
| Age (yrs) | 54.56 (16.00) | 43.45 (14.29) | <0.001* |
| Step count | 6,990 (2,540) | 5,580 (2,050) | <0.001* |
| Sedentary minutes | 692.75 (91.21) | 710.89 (81.08) | 0.123 |
| Total sleep minutes | 400.52 (47.59) | 403.10 (52.33) | 0.716 |
| Deep sleep minutes | 59.85 (15.05) | 64.69 (15.33) | 0.024* |
| REM sleep minutes | 80.93 (19.72) | 83.90 (18.57) | 0.260 |
| Deep sleep % | 15.01 (3.60) | 16.17 (3.66) | 0.023* |
| REM sleep % | 19.95 (4.18) | 20.45 (3.34) | 0.323 |
| Awake time % | 12.14 (1.68) | 12.46 (1.85) | 0.201 |
| Sex |  |  |  |
| Female | 127 (67.2%) | 62 (32.8%) | — |
| Male | 47 (79.7%) | 12 (20.3%) | — |
| Race |  |  |  |
| White | 122 (66.3%) | 62 (33.7%) | — |
| Non-White | 52 (81.2%) | 12 (18.8%) | — |

The sample was predominantly female. Based on row percentages in Table 1, 32.8% of females and 20.3% of males screened positive for moderate-to-severe depressive symptoms. Differences across racial groups were also observed, with 33.7% of White participants and 18.8% of non-White participants screening positive.

Several demographic and behavioral differences were observed between groups based on two-sample t-tests of subject-level measures. Participants with moderate-to-severe depressive symptoms were younger on average than those with none-to-mild symptoms (43.45 vs. 54.56 years) and had lower average daily step counts (5,580 vs. 6,990 steps), with both differences reaching statistical significance (Table 1).

Sleep duration and sedentary time were similar across groups, with comparable total sleep minutes (403.10 vs. 400.52 minutes) and sedentary time (710.89 vs. 692.75 minutes). In contrast, modest but statistically significant differences were observed in deep sleep duration, with participants in the moderate-to-severe group spending more time in deep sleep on average (64.69 vs. 59.85 minutes), corresponding to a slightly higher proportion of total sleep time (16.17% vs. 15.01%). Other sleep measures, including REM sleep duration (83.90 vs. 80.93 minutes), REM sleep percentage (20.45% vs. 19.95%), and awake time percentage (12.46% vs. 12.14%), were similar across groups.

These descriptive comparisons reflect unadjusted differences based on subject-level summaries and should be interpreted independently of the multivariable regression results presented below. In particular, the observed difference in deep sleep duration was not statistically significant after adjustment for demographic and behavioral covariates, and this variable was therefore not retained in the primary logistic regression model.

Figure 2 illustrates the distributions of age, average steps, sleep characteristics, and step trends over time, stratified by depressive symptom severity. Participants with moderate-to-severe depressive symptoms tended to be younger, with a leftward shift in the density relative to those with none-to-mild symptoms. A similar pattern was observed for average daily steps, reflecting lower physical activity levels among those with higher depression severity.

**Figure 2.**
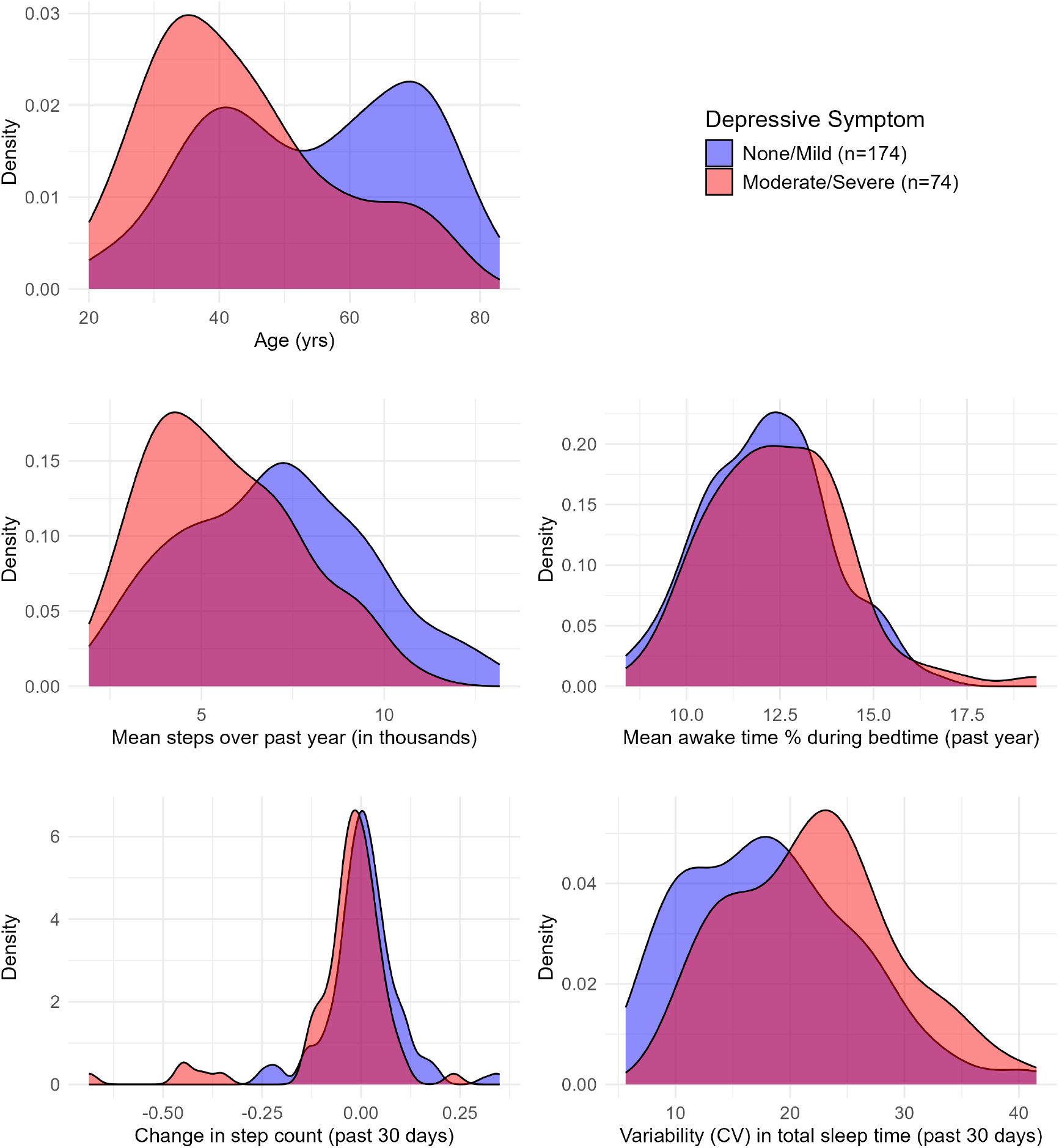
Distributions of continuous variables stratified by depressive symptom severity.

The distributions for average awake time percentage after sleep onset were largely overlapping between groups, suggesting minimal differences in sleep fragmentation. In contrast, variability in sleep duration over the past 30 days showed a rightward shift in the moderate-to-severe group, indicating greater night-to-night inconsistency.

The change in step count over the past 30 days also differed between groups. Among participants with none-to-mild symptoms, the distribution was centered near zero, indicating relatively stable step counts. In contrast, the distribution for the moderate-to-severe group was centered below zero, suggesting a downward trend in activity over time.

### 3.2 Exploratory longitudinal trends

To visualize temporal patterns in activity and sleep across depressive symptom severity, we examined marginal trends without covariate adjustment. Figure 3 shows smoothed trajectories of daily step counts and awake time percentage after sleep onset using generalized additive models (GAMs), which allow flexible modeling of nonlinear relationships (Hastie and Tibshirani, 1986; Wood, 2017). These exploratory models were used for visualization only and were not part of the primary inferential analyses.

**Figure 3.**
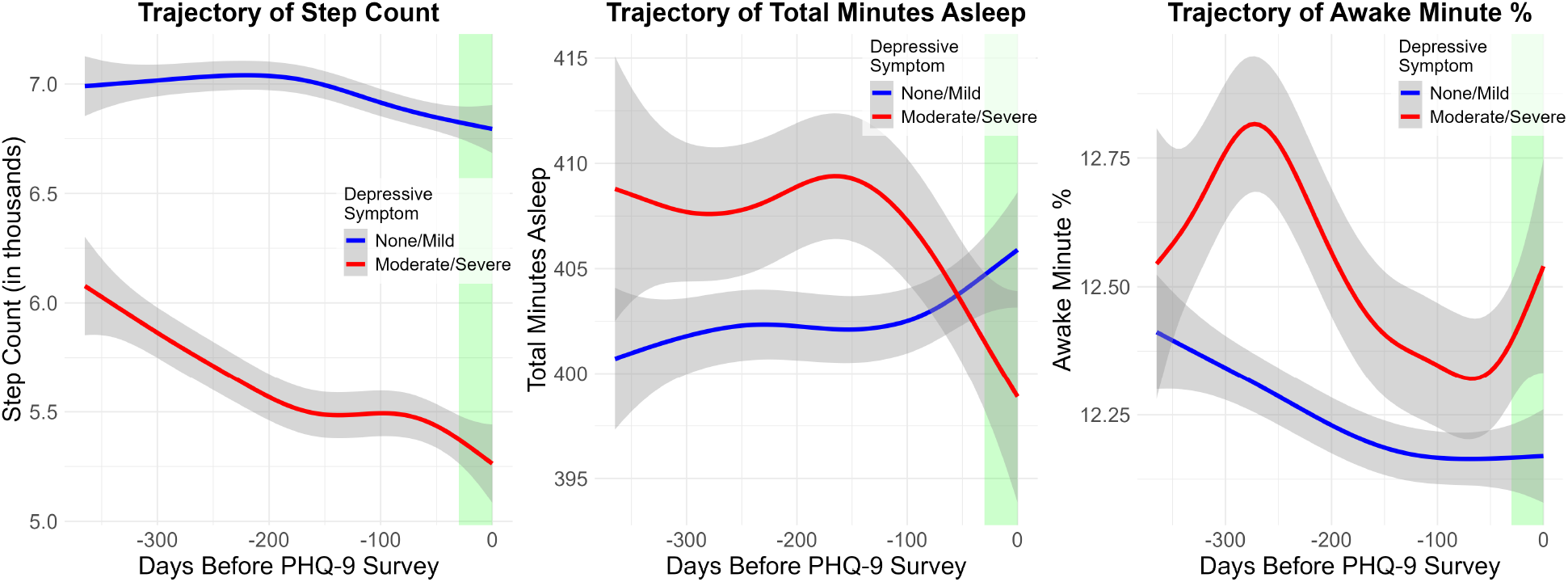
Smoothed GAM trajectories of step count, total sleep duration, and awake time percentage over the past year relative to PHQ-9 assessment. Curves represent group-level averages stratified by depressive symptom severity. Shaded bands indicate 95% confidence intervals. Green regions highlight the 30-day window prior to assessment.

Participants with moderate-to-severe depressive symptoms exhibited lower and gradually declining step-count trajectories over the observed period, whereas those with none-to-mild symptoms maintained higher and more stable activity levels. In contrast, awake time percentage after sleep onset exhibited greater variability in the moderate-to-severe group.

### 3.3 Logistic regression results

Table 2 presents logistic regression results examining associations between wearable-derived behavioral measures and the odds of moderate-to-severe depressive symptoms (PHQ-9 ≥ 10), adjusting for age, sex, and race. Model diagnostics indicated no evidence of multicollinearity, with all variance inflation factors below 2.

**Table 2:**
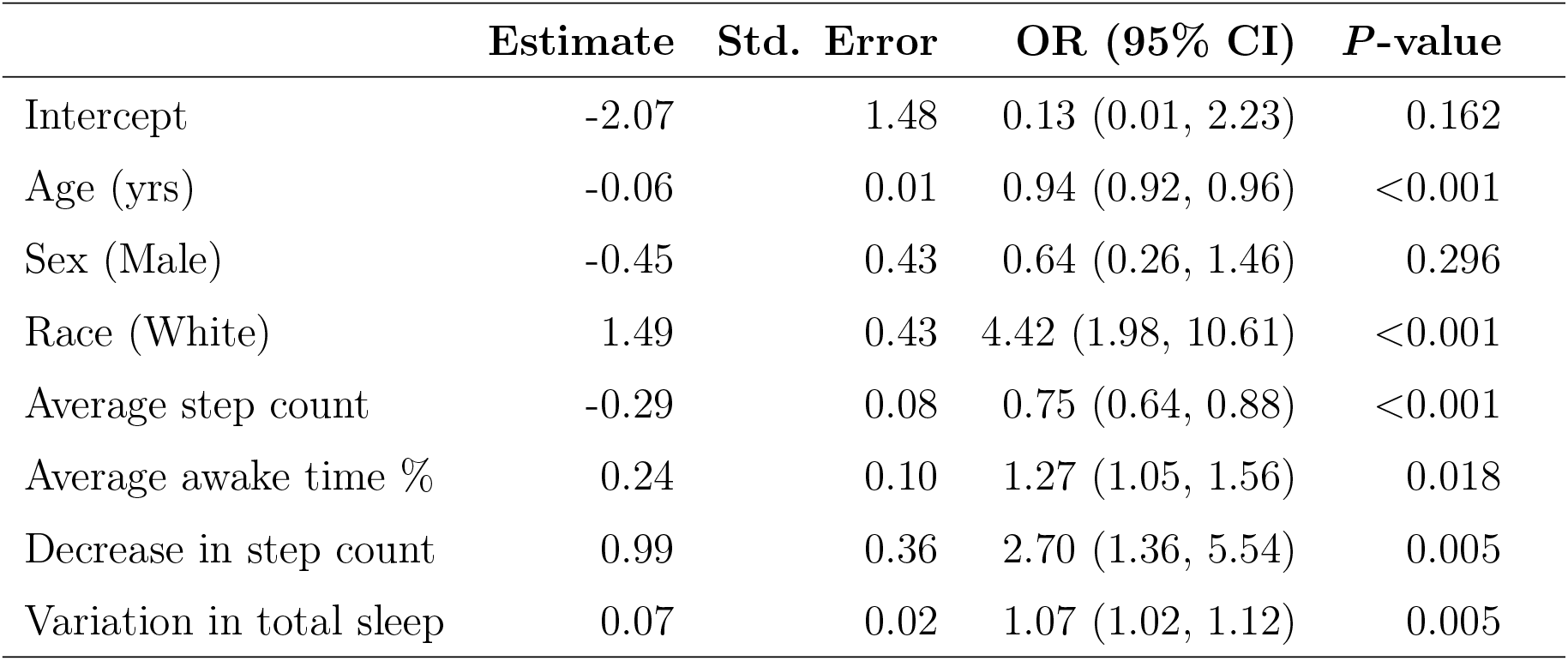
Logistic regression results predicting moderate-to-severe depressive symptoms using Fitbit-derived long-term and short-term features. Model adjusted for demographics. OR = odds ratio, CI = confidence interval.

Older age was associated with lower odds of moderate-to-severe depressive symptoms (OR = 0.94, 95% CI: 0.92–0.96; *p* < 0.001), corresponding to a 6% decrease in odds per year. Within this analytic sample, White participants demonstrated higher observed odds of moderate-to-severe depressive symptoms relative to the aggregated non-White category (OR = 4.42, 95% CI: 1.98–10.61; p < 0.001). This direction is counterintuitive relative to the broader epidemiological literature and likely reflects sample-specific factors, including differential self-report of depressive symptoms, selection into wearable device use, and the heterogeneity masked by the White versus non-White collapse. This finding should be interpreted cautiously, and race is included here as a demographic control variable rather than a variable of primary interest. Sex was not significantly associated with depressive symptom status.

Higher long-term physical activity was associated with lower odds of depressive symptoms. Each additional 1,000 steps per day over the past year was associated with a 25% reduction in odds (OR = 0.75, 95% CI: 0.64–0.88; *p* < 0.001), suggesting that higher habitual activity levels were associated with lower depressive symptom severity. In contrast, each 1% increase in average awake time after sleep onset was associated with a 27% increase in odds (OR = 1.27, 95% CI: 1.05–1.56; *p* = 0.018), suggesting that greater sleep fragmentation may be linked to elevated depressive symptoms.

Short-term behavioral dynamics were also associated with depressive symptoms. A binary indicator of declining step count over the past 30 days was associated with a 170% increase in odds (OR = 2.70, 95% CI: 1.36–5.54; *p* = 0.005), suggesting that recent decreases in activity may reflect behavioral changes linked to depressive symptom severity. Greater variability in sleep duration was similarly associated with increased odds, with each 1 percentage point increase in variability corresponding to a 7% increase in odds (OR = 1.07, 95% CI: 1.02–1.12; *p* = 0.005), suggesting that irregular sleep patterns may also reflect elevated depressive symptom severity.

Predictive performance comparisons are shown in Figure 4. The demographics-only model achieved an average AUC of 0.73. Adding short-term behavioral measures yielded an average AUC of 0.77, while adding long-term behavioral measures alone yielded an average AUC of 0.77. The combined model achieved the highest average AUC of 0.80, suggesting that long-term behavioral patterns and short-term behavioral changes provide complementary information about depressive symptom severity.

**Figure 4.**
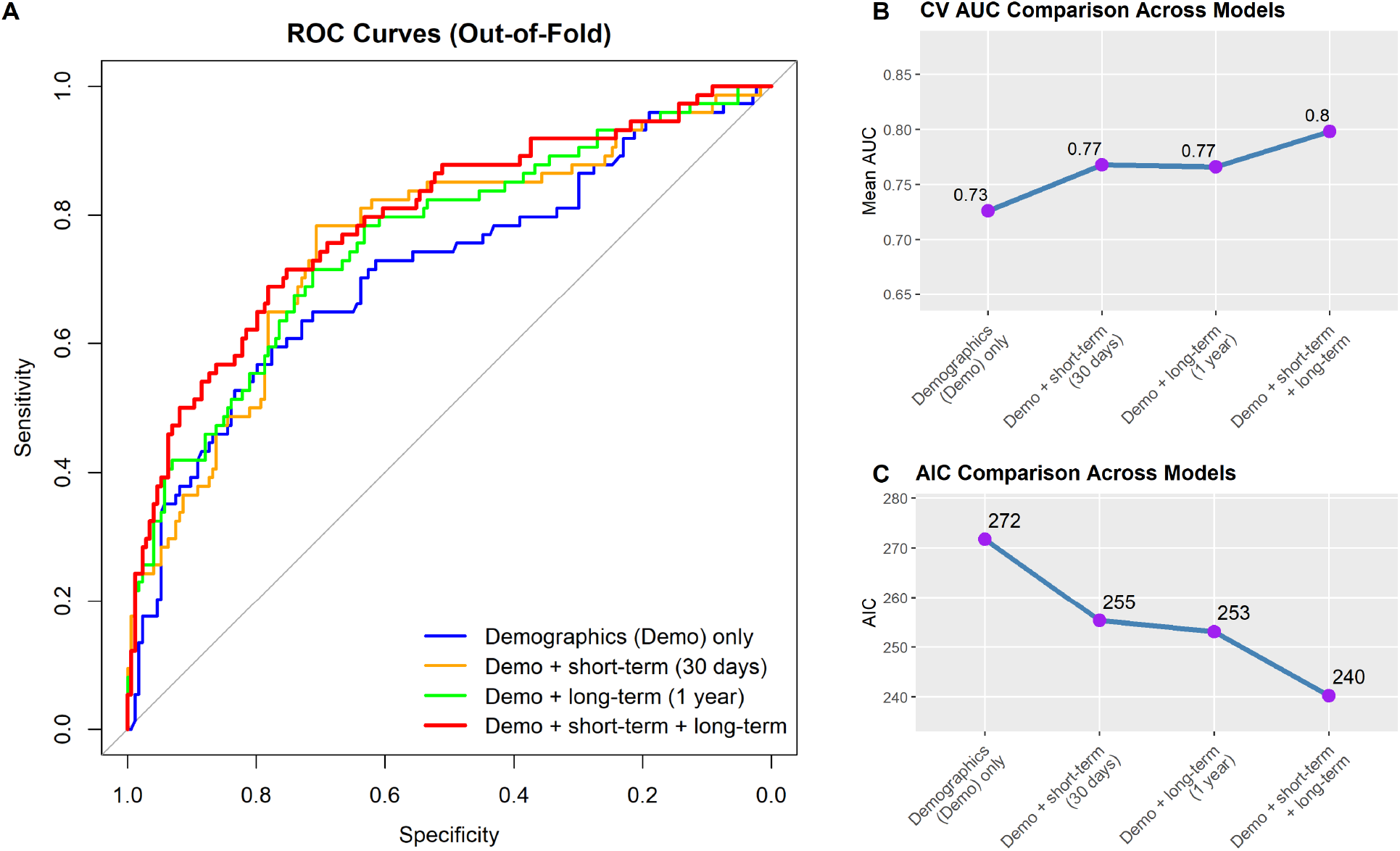
Model performance comparisons. (A) ROC curves. (B) AUC values. (C) AIC values across models.

Model fit improved across models, with AIC decreasing from 271.7 for the demographics-only model to 240.3 for the combined model, indicating improved overall fit with the inclusion of wearable-derived behavioral measures.

Sensitivity analyses using continuous PHQ-9 scores produced directionally consistent findings. Lower long-term step count, greater awake time after sleep onset, declining short-term step trajectories, and greater sleep variability remained associated with greater depressive symptom severity, suggesting that the observed associations were not solely dependent on dichotomization of the PHQ-9 outcome. Results were also directionally consistent when the binary indicator of declining step count was replaced with the continuous step-count slope estimate, indicating that associations with depressive symptom severity were robust to alternative parameterizations of short-term step trajectories.

## 4 Discussion

In this study, we examined associations between wearable-derived behavioral measures and depressive symptom severity in a sample of All of Us participants, finding that both long-term behavioral patterns and short-term changes in physical activity and sleep were associated with moderate-to-severe depressive symptoms beyond demographic characteristics alone. These findings support the potential utility of passively collected wearable data for characterizing behavioral patterns associated with depressive symptom severity in naturalistic settings.

A central contribution of this study is the explicit distinction between long-term habitual behavioral patterns and short-term behavioral dynamics preceding PHQ-9 assessment. Whereas many prior wearable-based studies have primarily relied on static summaries averaged across broad time windows, our framework evaluates whether recent within-person behavioral changes provide additional information beyond chronic baseline behavior. The observed improvement in model discrimination when combining both temporal scales suggests that behavioral trajectories may capture dynamic aspects of depressive symptom severity not fully reflected by long-term averages alone. In addition, aligning wearable-derived measures relative to each participant’s PHQ-9 survey date enabled consistent comparison of behavioral windows across individuals, while the use of repeated stratified cross-validation provided a more stable estimate of model performance than a single train-test split (Burman, 1989).

Higher habitual step count was associated with lower odds of moderate-to-severe depressive symptoms (OR = 0.75 per 1,000 steps/day), consistent with prior literature linking physical activity to depression risk and symptom severity (Bizzozero-Peroni et al., 2024; Pearce et al., 2022). Beyond average activity levels, a declining trend in step count over the 30 days preceding assessment was independently associated with increased odds of depressive symptoms (OR = 2.70). These findings suggest that short-term behavioral trajectories may capture clinically relevant changes beyond stable activity levels alone. Declining physical activity may reflect reduced motivation, fatigue, psychomotor slowing, and withdrawal from daily activities commonly associated with depression (American Psychiatric Association, 2013). Prior studies have similarly demonstrated that within-person decreases in activity are associated with worsening depressive symptoms (Snippe et al., 2016; Lindwall et al., 2014). Our findings extend this literature by suggesting that recent changes in activity captured through wearable devices may provide passively measured behavioral indicators associated with elevated depressive symptom severity.

Sleep-related findings further highlight the importance of behavioral dynamics. Higher average awake time percentage after sleep onset was associated with increased odds of depressive symptoms (OR = 1.27), consistent with evidence linking sleep fragmentation to depression (Maki et al., 2025; Baglioni et al., 2011). Greater variability in sleep duration over the 30 days preceding assessment was also associated with elevated depressive symptoms (OR = 1.07). The use of the coefficient of variation to quantify sleep irregularity represents a methodological strength because it normalizes variability relative to individual baseline sleep duration, facilitating comparison across participants with different average sleep durations. These results align with prior work suggesting that irregular sleep patterns are associated with adverse mental health outcomes (Lim et al., 2022; Lunsford-Avery et al., 2018, 2022), and further support the importance of sleep regularity beyond duration alone. Irregular sleep patterns may reflect disruptions in circadian and behavioral regulation associated with impaired emotional functioning and depressive symptoms.

Model performance results suggest that jointly incorporating both short- and long-term behavioral measures yielded the strongest model discrimination. The combined model achieved the highest cross-validated average AUC (0.80), compared to 0.77 for short-term measures alone, 0.77 for long-term measures alone, and 0.73 for the demographics-only baseline. Because repeated stratified cross-validation was used, these AUC estimates reflect out-of-sample performance rather than in-sample fit, which partially mitigates concerns about overfitting given the modest EPV. Together, these findings suggest that long-term behavioral patterns and recent behavioral changes provide complementary information regarding depressive symptom severity. The observed decrease in AIC from 271.7 for the demographics-only model to 240.3 for the combined model further suggests that wearable-derived behavioral measures contribute nonredundant information beyond demographic characteristics alone. Importantly, these models were not intended for clinical diagnosis or screening deployment. Rather, the findings demonstrate associations between wearable-derived behavioral patterns and depressive symptom severity within a research setting. Additional prospective validation in larger and more representative cohorts would be necessary before considering clinical implementation. The observed discrimination should therefore be interpreted as exploratory evidence supporting the potential utility of wearable-derived behavioral measures as adjunctive behavioral markers associated with depressive symptom severity.

Our findings are consistent with prior studies reporting moderate-to-good classification performance using wearable- and smartphone-derived behavioral measures (Moshe et al., 2021; Rohani et al., 2018). This growing body of literature suggests that passive behavioral sensing may provide scalable approaches for monitoring behavioral correlates of depressive symptom severity in real-world settings.

With respect to demographic factors, older age was associated with lower odds of depressive symptoms, consistent with epidemiological trends (Lee et al., 2023). Race was included as a covariate to adjust for demographic differences in depressive symptom reporting and wearable usage patterns and should not be interpreted as a primary finding of this study. Within this analytic sample, White participants demonstrated higher observed odds of moderate-to-severe depressive symptoms relative to the aggregated non-White category (OR = 4.42), a direction that differs from broader epidemiological literature documenting substantial depression burden and unmet treatment need across many non-White populations (Williams and Mohammed, 2009; Gonzalez et al., 2010). Several sample-specific factors may contribute to this pattern. Differences in depressive symptom reporting may reflect cultural, structural, or healthcare-access factors that influence how symptoms are recognized, reported, or clinically identified across populations (Gonzalez et al., 2010). In addition, individuals who actively use wearable devices and participate in a research cohort such as All of Us may represent a more health-engaged subset of the population, potentially introducing selection bias. The aggregation of heterogeneous racial and ethnic groups into a single non-White category, which was necessary because of limited subgroup sample sizes, may further obscure meaningful heterogeneity across groups. Consequently, this finding should be interpreted cautiously and not generalized beyond the present sample. More broadly, observed racial differences in wearable-derived behavioral measures and depressive symptom reporting likely reflect the influence of structural determinants of health, including socioeconomic conditions, healthcare access, occupational demands, stress exposure, and technology engagement, rather than intrinsic biological differences. Future studies with larger and more diverse cohorts are needed to evaluate behavioral dynamics across more granular racial and ethnic subgroups and to better understand how these structural and social determinants interact with wearable-derived behavioral markers.

Future work should also evaluate whether these behavioral dynamics prospectively predict changes in depressive symptom severity over time and whether wearable-derived markers can be meaningfully integrated into clinical monitoring workflows for depression management.

Several limitations should be considered. The relatively small sample size (*N* = 248, with 74 outcome-positive participants) may affect the stability of coefficient estimates and model discrimination, and the EPV is near the conventional threshold of 10. Although repeated cross-validation provides more stable AUC estimates than a single train-test split, results should be treated as exploratory and replicated in larger cohorts before drawing firm conclusions. Participants contributing longitudinal Fitbit data may differ systematically from the broader population with respect to socioeconomic status, digital literacy, healthcare engagement, and health behaviors. Individuals who consistently wear and synchronize wearable devices may also exhibit healthier baseline behaviors or greater interest in self-monitoring, potentially limiting generalizability. Participants with more severe depressive symptoms may also demonstrate lower wearable adherence, potentially introducing informative missingness that could bias behavioral summaries. Wearable-derived measures are additionally subject to device-specific measurement error, variable wear adherence, and missingness that may not occur completely at random. The cross-sectional nature of the PHQ-9 assessment further limits causal interpretation. Because depressive symptoms and wearable-derived behaviors may influence one another bidirectionally, the present findings should not be interpreted causally. Although demographic covariates were included, residual confounding related to socioeconomic conditions, occupational schedules, chronic illness burden, medication use, and lifestyle factors may remain. Finally, the aggregation of heterogeneous racial and ethnic groups into a single non-White category was performed solely to reduce sparse-data instability and should not be interpreted as reflecting homogeneous behavioral or mental health patterns across groups. The observed direction of the race association likely reflects sample-specific factors, including differential symptom reporting, selection into wearable device use, and the collapse of diverse racial and ethnic groups into a single analytic category, rather than true population-level differences. Larger studies with sufficient subgroup representation are needed to evaluate potential heterogeneity across more granular racial and ethnic categories.

Despite these limitations, this study highlights the potential of wearable-derived behavioral data for characterizing patterns associated with depressive symptom severity. In particular, recent changes in activity and sleep may provide behavioral signatures of elevated depressive symptoms beyond long-term habitual behavioral measures alone. Because wearable devices continuously capture behavior in real-world settings, these approaches may support future efforts aimed at passive behavioral monitoring and the identification of behavioral patterns associated with worsening depressive symptom severity between clinical encounters. Clinically, this may be particularly relevant for individuals who are seen infrequently or who may underreport symptom changes, for whom objective behavioral measures could complement traditional self-report assessments. Future work integrating wearable-derived markers into clinical workflows warrants further investigation, including consideration of both the potential utility and ethical implications of passive behavioral monitoring in mental health care.

## CRediT Authorship Contribution Statement

**Jenifer Rim**: Writing – Original Draft, Visualization, Methodology, Formal analysis, Conceptualization, Software. **Qi Xu**: Conceptualization, Methodology, Writing – Review & Editing. **Xiwei Tang**: Conceptualization, Methodology, Writing – Review & Editing, Funding acquisition. **Cadence Pinkerton**: Writing – Original Draft, Project administration. **Yuqing Guo**: Conceptualization, Writing – Review & Editing, Supervision, Funding acquisition. **Annie Qu**: Conceptualization, Methodology, Writing – Review & Editing, Supervision, Project administration, Funding acquisition.

## Ethics Approval and Consent to Participate

This study used de-identified data from the All of Us Research Program (Registered Tier Dataset v8). The All of Us Research Program is overseen by an Institutional Review Board (IRB) and all participants provided informed consent prior to enrollment. This secondary analysis of publicly available, de-identified data was conducted in accordance with the All of Us Data Use Agreement and did not require additional IRB review.

## Data Availability Statement

The data used in this study are available through the All of Us Research Program Researcher Workbench (https://workbench.researchallofus.org). Access requires registration and approval through the All of Us Research Program (https://www.researchallofus.org).

## Acknowledgments

This research was supported by National Cancer Institute SCH: Individualized learning and prediction for heterogeneous multimodal data from wearable devices (R01: 1R01CA297869 to AQ and YG), National Science Foundation Data integration for heterogeneous data: A general framework for distribution shift, posterior drift and block missing data (DMS: 2515275 to AQ), and NSF grant: High-Dimensional Point Process Modeling with Applications to Large-scale Neuronal Activity Analysis (DMS: 2113467 to XT).

